# EMS injury cause codes more accurate than emergency department visit ICD-10-CM codes for firearm injury intent in North Carolina

**DOI:** 10.1101/2023.11.21.23298854

**Authors:** Nicole L. Snyder, Amy Ising, Anna E. Waller

**Author notes:** Corresponding author (AI).

## Abstract

**Background:** The timeliness, accuracy, and completeness of data for firearm injury surveillance is crucial for public health surveillance efforts and informing injury prevention measures. While emergency department (ED) visit data can provide near real-time information on firearms injuries, there are concerns surrounding the accuracy of intent coding in these data. We examined whether emergency medical service (EMS) data provide more accurate firearm injury intent coding in comparison to ED data.

**Methods:** We applied a firearm injury definition to EMS encounter data in NC’s statewide syndromic surveillance system (NC DETECT), from January 1, 2021, through December 31, 2022. Each record was manually reviewed to determine shooter and intent, and the corresponding manual classifications were compared to the injury cause codes entered in the EMS data and to ED visit records, where linkage was possible.

**Results:** We identified 9557 EMS encounters from January 1, 2021, through December 31, 2022 meeting our firearm injury definition. After removing false positives and duplicates, 8584 records were available for manual injury classification. Overall, our analysis demonstrated that manual and EMS injury cause code classification were comparable. However, for the 3401 EMS encounters that could be linked to an ED visit record, only 18.3% (n = 355) of the 1945 assaults and 22.2% (n = 38) of the 171 intentional self-harm suicide encounters we identified in the EMS records were identified as assault firearm injures in the ED visit data. This demonstrates a marked difference in the intent coding between the two data sources.

**Conclusions:** This study illustrates both the value of examining EMS encounters for firearm injury intent, and the challenges of accurate intent coding in the ED setting. The results also suggest the importance of developing better guidance around intent coding for firearm injuries in the ED.

## Introduction

As firearm injury continues to be a significant cause of morbidity and mortality in the United States (US), public health researchers continue to seek improvements to the timeliness, accuracy, and completeness of firearm injury surveillance [1]. In North Carolina (NC), firearm injury surveillance relies primarily on the NC Violent Death Reporting System (NC-VDRS), hospital administrative data, and syndromic surveillance emergency department visit data [2]. While the syndromic surveillance emergency department (ED) visit data provides near real time information on nonfatal firearm injury in NC, there are concerns about accuracy when subtyping firearm injuries by intent (assault, unintentional, intentional self-harm). For example, while some studies estimate unintentional firearm injury to represent 16% or less of all nonfatal firearm injury, approximately 60% of ED firearm visits in NC are identified as unintentional [3, 4]. Several other studies that have documented these coding intent discrepancies and the potential overclassification of nonfatal firearm injury as unintentional, relied on urban, Level I Trauma center data from a small number of hospitals [5, 6]. For comparison, we aimed to quantify any coding intent discrepancies using data from a statewide system in a predominantly rural state. To achieve this, we compared firearm injury intent classifications as identified in statewide emergency medical services (EMS) encounter data to the firearm injury intent classifications identified in syndromic surveillance ED visit data, for those patients transported to the ED by EMS. We specifically chose to examine EMS data because EMS narratives provide rich contextual details on the entire EMS encounter and can, therefore, provide insights into firearm injury intent.

## Methods

We conducted this evaluation in several steps. First, we applied a firearm injury definition to EMS encounter data for encounters sent to NC DETECT, NC’s statewide syndromic surveillance system, from January 1, 2021 through December 31, 2022. We then manually reviewed each record and documented shooter and intent. Third, we compared manual intent classifications to those documented in the EMS data based solely on injury cause code. Finally, we linked the EMS records to corresponding ED visit records, where possible, to compare our manually assigned intent classifications to the intent coding in the ED visit data. The UNC IRB determined this study to be exempt from full review.

### EMS firearm V1 definition

The EMS firearm definition we used for this analysis is outlined in Table 1. It searches the complaint for gunshot wound related injuries OR the injury cause field for firearm-related ICD-10-CM codes. The definition also searches for gunshot wound related injuries in the narrative if the dispatch complaint is *Stab/Gunshot Wound/Penetrating Trauma*. This initial definition was designed to maximize sensitivity, while excluding EMS encounters that are not for active responses based on EMS patient disposition.

**Table 1.** EMS Firearm V1 Definition.

|  |  |
| --- | --- |
| Complaint Terms | GSW or gunshot or "gun shot" or "shot myself" or "shot self*" or firearm or "I got shot" or "been shot" or "shot gun" or shotgun |
| Injury cause codes | W34, W34.0, W34.09, W34.00, W34.00XA, W33.02, W33.03, W33.09, W33.01, W32.0, W32, W34.1, W33.0, W33, X94.0XXA, X95.9, X95.9XXA, Y22, Y22.XXXA, Y23.1, X72.XXXA, X74.9, Y35.0, Y35.02, Y35.023, Y35.03, Y35.092, Y35.093, Y35.003, Y23.3, Y22.0, Y24.8, Y23.8, Y23, Y23.0, Y23.0XXA, Y38.4X, Y24.9, Y24.9XXA, Y23.9 |
| Dispatch complaint AND narrative | dispatch complaint is Stab/Gunshot Wound/Penetrating Trauma AND narrative keywords of GSW or gun shot or gunshot |
| Patient disposition exclusions | Assist, Agency; Assist, Public, Assist; Unit, Canceled (Prior to Arrival At Scene); Canceled on Scene (No Patient Contact); Canceled on Scene (No Patient Found); Standby-No Services or Support Provided; Standby-Public Safety, Fire, or EMS Operational Support Provided; Transport Non-Patient, Organs, etc. |

### EMS encounter manual review

With approval from the NC Office of Emergency Medical Services, we downloaded all EMS encounters from January 1, 2021 through December 31, 2022 meeting the firearm v1 definition from NC’s statewide syndromic surveillance system NC DETECT on February 1, 2023 with an update to incorporate additional records on May 1, 2023[7]. The authors did not have access to any information that could identify individual participants during or after data collection. For each EMS encounter, we manually documented the shooter (self, other, unknown) and intent (intentional, unintentional, undetermined) in a spreadsheet, which we also collapsed into the categories used in injury intent coding in the International Classification of Diseases: assault (other – intentional), self-inflicted (self – intentional), unintentional (other, unknown, and self) and undetermined (other, unknown, and self). We also indicated false positives. We then added notes, as needed, to document our rationale for any of these classifications. EMS encounters were reviewed by only one person, but all challenging EMS encounters were reviewed by three reviewers to achieve consensus on coding. We included both non-fatal and fatal firearm injuries in our review. Additional decision points are listed below:

- Intent was assigned using complaint, narrative, and injury cause code fields. Sometimes these fields provided conflicting information. When that occurred, the reviewer used group consensus to determine intent.
- If injury cause was coded as undetermined, a more precise intent may have been assigned based on the narrative description.
- When the narrative described an incident as a drive-by shooting or shot by someone else while in a car with no documented/undetermined injury cause, intent was classified as assault.
- If a patient was transported to an ED by EMS, stabilized, and then transferred by EMS to another ED, both EMS encounters were coded.
- For deaths at the scene with inconclusive intent in the EMS record, death certificate data were consulted to determine intent, if a linkage could be identified based on patient age, sex, race, county of residence and date of death.

### False positives

We excluded any encounters identified by the firearm v.1 definition that described injuries caused by bb guns, pellet guns, nail guns, air guns, or firearms used as a blunt weapon, e.g., “pistol whip.” We also excluded subsequent encounters for previous firearm injury and near misses.

### EMS-specific descriptive analyses

After we completed the manual review and removed false positives, we conducted a descriptive analysis of EMS firearm encounters. We calculated counts and crude rates statewide and by EMS county by sex, race, ethnicity, and age group. We compared the intent as identified in the injury cause field to the intent assigned manually during the review process. We classified EMS encounters by injury cause code into assault, intentional self-harm/suicide, unintentional, undetermined, and a final category of non-firearm injury cause codes. If the EMS encounter record included two conflicting firearm injury cause codes, we classified the encounter based on the first code listed.

### EMS-ED linkage

The EMS encounters from January 1, 2021 through December 31, 2022 meeting the EMS firearm v1 definition were linked, when possible, to the corresponding ED visit record and provided to the authors for analysis on October 19, 2023. The linkage used an existing hierarchical deterministic process to match records based on drop-off time by EMS, arrival time in the ED, EMS destination name, EMS destination type, patient date of birth, sex and race/ethnicity, ED facility name, and ED transport mode [8]. The linkage rules use the same variables across multiple steps with different matching criteria to maximize the number of linkages while minimizing false linkages. Several updates were made to the linkage process as part of this analysis to increase the likelihood of a successful linkage; we updated the EMS encounter destination type required for linkage to include missing and *Freestanding Emergency Department* destination types as well as *Hospital-Emergency Department*. We also allowed records in the linkage process with missing race and updated the free text list of hospital names used by EMS agencies in the Destination Name field to include additional aliases and misspellings, increasing that list from 3213 to 5739 entries.

### EMS-ED linkage descriptive analyses

We determined the number and percentage of linked EMS records to corresponding ED visits and compared the EMS manually assigned intent to the intent in the ED visit data as identified by ED firearm injury intent definitions developed by the Centers for Disease Control and Prevention (CDC) and implemented in NC DETECT [9, 10].

## Results and discussion

We identified 9557 EMS encounters from January 1, 2021, through December 31, 2022 meeting our firearm injury definition. Of these, 969 (10%) were identified as false positives and four records were duplicates, leaving 8584 EMS encounters for this analysis.

### False positive analysis

We grouped the 969 false positives we observed in our data into seven categories, as shown in Table 2. We then analyzed the false positives to inform improvements to our EMS firearm injury definition.

**Table 2.** EMS Syndrome Definition False Positive Categories.

| False Positive Category | Count (%) |
| --- | --- |
| Other types of injuries or other types of false positives that could not be categorized | 222 (22.9%) |
| Pistol whips with no penetrating firearm discharge injury | 185 (19.1%) |
| Subsequent encounters for a previous firearm injury | 173 (17.9%) |
| Firearm injuries by bb guns, pellet guns, air guns, nail guns, etc. | 121 (12.5%) |
| Near misses, e.g., injuries sustained near gunshots but with no evidence of direct injury by a bullet | 113 (11.7%) |
| Mental health concerns without firearm injury | 92 (9.5%) |
| Stabbings | 63 (6.5%) |

The largest number of false positives, 22.9% (n = 222) fell into the *other types of injuries* category, which included EMS encounters with no documented firearm discharge and other types of assault. Pistol whips were also a common source of false positives; 173 (93.5%) of these had a firearm injury cause code, typically *Assault by unspecified firearm discharge (X95.9)*, even though there was no documented firearm discharge injury in the EMS record. We often identified subsequent encounters (n = 173, 17.9%) in the EMS narrative text; 80 (46.2%) of these subsequent encounter records had a firearm discharge injury cause code. Injuries sustained near gunshots but with no evidence of direct injury by a bullet, e.g., injuries sustained by falls running away from gunshots, lacerations sustained from broken glass caused by bullets hitting windshields and windows, etc., comprised 11.7% (n = 113) of false positives. Examples of the 92 encounters (9.5%) that fell into the mental health category included family members of firearm injury victims who had a mental health crisis after the firearm injury event, suicidal patients with firearms who did not suffer firearm injury, patients with mental health conditions claiming firearm injury with no identified firearm injury, and patients with substance misuse claiming firearm injury with no identified firearm injury. The 63 (6.5%) stabbing-related false positives resulted primarily from including the *Stab/Gunshot Wound/Penetrating Trauma* dispatch complaint in our definition and this dispatch complaint text being repeated in the narrative text. Based on our false positive analysis, we hypothesize that the addition of chief complaint exclusion terms used in the CDC-developed ED firearm definition may improve our EMS firearm definition [9]. Additionally, exclusion terms that address false positives, e.g., pistol whips, even when the encounter has a firearm discharge injury cause code, will require logic that applies these exclusion terms to encounters with firearm discharge injury cause codes in addition to exclusion terms specific to the complaint and narrative free text fields.

### EMS descriptive analyses

The primary cause of firearm related injury in 2021-2022 EMS encounter data was assault, which encompassed 51.8% (n = 4446) of all recorded events, followed by undetermined (n = 1694; 19.7%), intentional self-harm (n = 1581;18.4%), and unintentional (n = 862; 10.1%). The ICD-10-CM injury intent coding does not classify unintentional and undetermined firearm injury by perpetrator, but our manual intent coding revealed that 7.3% (n = 634) of EMS firearm encounters were for unintentional self-inflicted injury, e.g., persons shooting themselves while cleaning, holstering, storing, retrieving, stepping on and/or dropping firearms. This finding presents an opportunity to inform unintentional firearm injury prevention messaging with specific scenarios. For example, public health messaging could remind firearm owners to clean their firearms while sober, and to store their firearms unloaded.

### EMS demographic analysis

We categorized firearm injury intent by key demographics, revealing several important differences by sex, age group, and race/ethnicity, as shown in Table 3. Notably, EMS firearm injuries were more than 7.0 times more likely to occur among males in comparison to females. Males were also 4.7 times more likely to be involved in an assault-related firearm injury and 6.2 times more likely to be involved in an intentional self-inflicted firearm injury compared to females. With respect to age group, the highest rate of firearms related assaults was among those aged 19 to 24 (112.77 events per 100,000), followed by those aged 15 to 18 (92.90 events per 100,000). In contrast, those aged 65+, followed closely by those aged 19 to 24 years of age, were more likely to be involved in an intentional self-inflicted firearm injury, with rates of 23.31 per 100,000 and 19.28 per 100,000, respectively. For race/ethnicity, Black or African Americans were 13.3 times more likely to be involved in a firearm injury related to assault in comparison to White persons, while White persons had the highest rate of intentional self-inflicted firearm injury, highlighting the disparities among these two demographic groups. Notably, American Indian/Alaskan Native (AIAN) persons had the second highest assault firearm injury rate, which was nearly four times higher than Whites, at 41.43 per 100,000.

**Table 3.** EMS Firearm Injury Demographics, by Count and Crude Rate.^a^

| **Demographic** | **Assault** | **Intentional**<br>**Self-Harm** | **Undetermined** | **Unintentional** | **All**<br>**Intents** |
| --- | --- | --- | --- | --- | --- |
| Total | 4446<br>(39.17) | 1581<br>(13.93) | 1694<br>(14.93) | 863<br>(7.60) | 8584<br>(75.64) |
| **Sex** |  |  |  |  |  |
| Male | 3621<br>(70.34) | 1342<br>(26.07) | 1403<br>(27.25) | 743<br>(14.43) | 7109<br>(138.09) |
| Female | 808<br>(14.77) | 230<br>(4.21) | 270<br>(4.90) | 112<br>(2.05) | 1420<br>(25.99) |
| Unknown/Missing | 17 | 9 | 21 | 8 | 55 |
| **Age Group** |  |  |  |  |  |
| Infant/Preschool<br>(0-4) | 19<br>(3.20) | 0<br>(0.00) | 20<br>(3.37) | 26<br>(4.37) | 65<br>(10.93) |
| Elementary School<br>(5-9) | 23<br>(3.80) | 0<br>(0.00) | 19<br>(3.14) | 14<br>(2.31) | 56<br>(9.25) |
| Middle School<br>(10-14) | 86<br>(13.23) | 22<br>(3.38) | 36<br>(5.54) | 32<br>(4.92) | 176<br>(27.07) |
| High School<br>(15-18) | 515<br>(92.90) | 66<br>(11.91) | 199<br>(35.90) | 53<br>(9.56) | 833<br>(150.26) |
| College<br>(19-24) | 1012<br>(112.77) | 173<br>(19.28) | 354<br>(39.45) | 175<br>(19.50) | 1714<br>(190.99) |
| Young Adult<br>(25-44) | 2074<br>(75.25) | 434<br>(15.75) | 751<br>(27.25) | 311<br>(11.28) | 3579<br>(129.54) |
| Middle Aged<br>(45-64) | 580<br>(21.41) | 443<br>(16.35) | 229<br>(8.45) | 155<br>(5.72) | 1407<br>(51.93) |
| Senior<br>(65+) | 91<br>(4.93) | 430<br>(23.31) | 51<br>(2.76) | 90<br>(4.88) | 662<br>(35.88) |
| Unknown/Missing | 46 | 13 | 35 | 7 | 101 |
| **Race/Ethnicity** |  |  |  |  |  |
| American<br>Indian/Alaskan<br>Native | 74<br>(41.43) | 6<br>(3.36) | 61<br>(34.15) | 12<br>(6.72) | 153<br>(85.67) |
| Asian/Pacific<br>Islander | 22<br>(5.63) | 17<br>(4.35) | 7<br>(1.79) | 11<br>(2.82) | 57<br>(14.59) |
| Black or African<br>American | 3103<br>(139.28) | 211<br>(9.47) | 1020<br>(45.78) | 266<br>(11.94) | 4600<br>(206.47) |
| Hispanic or Latino | 281<br>(24.24) | 56<br>(4.83) | 89<br>(7.68) | 45<br>(3.88) | 471<br>(40.63) |
| White | 801<br>(10.83) | 1246<br>(16.85) | 351<br>(4.75) | 485<br>(6.56) | 2883<br>(38.99) |
| Unknown/Missing | 165 | 45 | 166 | 44 | 420 |

<sup>a</sup>Data are reported as count followed by rates per 100,000 persons within a given demographic in the state of North Carolina. For the total row, the total population was used for the denominator.

These results highlight the importance of targeted interventions. For example, holistic interventions that target middle-school and high-school age students could result in significant reductions in assault and intentional self-harm events. Recent efforts focused on establishing universal firearms risk injury screening in care settings for adolescents and youths [11], as well as developing a better understanding of firearm access among this group [12], may help drive preventative mechanisms in this area. Interventions that examine the underlying causes of the increasing rates of intentional self-harm among those aged 65+ [13], might lead to interventions such as increased health care provider communication and policies that address firearms access to those with signs of dementia and/or depression [14, 15]. Finally, holistic efforts to address the disparities that exist among different racial and ethnic groups with higher rates of firearm-related injury events [16] would be an important step forward for reducing injuries among these groups.

### Manual intent and injury cause code classification comparison

Next, we compared our manually assigned intent classifications to the EMS injury cause codes (ICD-10-CM) alone. The results are shown in Table 4. There were 2358 firearm injury encounters (27.5%) that were missing an injury cause code. Of the remaining 6226 EMS encounters, our manual intent coding was comparable to the intent identified solely by the injury cause code. Although our manual intent classifications referenced the injury cause codes during review, these results suggest that EMS injury cause codes can serve as a reliable indicator of firearm injury intent when available.

**Table 4.** Manual and Injury Cause Code Intent Classification Comparison, Count (Column %).

| Intent | Manual Injury Classification | EMS Injury Cause Code Classification* |
| --- | --- | --- |
| Assault | 4446 (51.8%) | 3055 (49.1%) |
| Intentional Self - Harm/Suicide | 1581 (18.4%) | 1111 (17.8%) |
| Unintentional | 862 (10.1%) | 580 (9.3%) |
| Undetermined | 1694 (19.7%) | 1211 (19.5%) |
| Multiple Intent Codes | --- | 105 (1.7%) |
| Other (non-firearm) | -- | 164 (2.6%) |
| Total | 8584 (100%) | 6226 (100%) |

\*Missing = 2,358

We further examined instances where our manual review did NOT match the intent as assigned by the EMS injury cause code. Our manual intent classifications did not agree with the assigned injury cause code intents for 773 EMS encounters, of which 641 (82.9%) were assigned an undetermined injury cause code. With manual review of the narrative text, we were able to assign 436 of these to assault, followed by 156 to intentional self-harm and 49 to unintentional. The manual intent assignments resulted in more specific intent coding rather than a change of intent for most EMS encounters. With an improvement in completeness, EMS injury cause codes alone can be a reliable source of intent-specific firearm injury surveillance.

### EMS disposition

We also grouped the EMS firearm injury encounters by EMS disposition to document both the continuum of care for patients initially encountered by EMS, and to assess the potential for linking each event to ED visit data (Table 5). Most events were recorded as ‘*Patient Treated, Transported by this EMS Uni*t’ (n = 6054, 71%). A total of 1889 patients (22%) were dead at the scene from firearm injury, 67% (n = 1062) of which were from intentionally self-inflicted gunshot wounds. For intents other than intentional self-harm, the majority of EMS encounters for firearm injury result in EMS transport.

**Table 5.** EMS Disposition by Intent by Count (Row %).

| Intent | All Encounters | Patient treated, transported by this EMS Unit | Patient Dead at Scene (with and without transport) | Other Disposition |
| --- | --- | --- | --- | --- |
| Assault | 4446 | 3484 (78%) | 602 (14%) | 360 (8%) |
| Intentional Self-Harm | 1581 | 467 (30%) | 1062 (67%) | 52 (3%) |
| Unintentional | 863 | 770 (89%) | 15 (2%) | 78 (9%) |
| Undetermined | 1694 | 1333 (79%) | 210 (12%) | 151 (9%) |
| All Intents | 8584 | 6054 (71%) | 1889 (22%) | 641 (7%) |

**Table 6.** Count (Row %) of EMS encounters linked to ED visits and binned into ED Firearm Syndromes by EMS Manual Intent.^a^

| Intent of EMS records assigned by manual review | EMS Firearm Encounters linked to ED visits | ED CDC Firearm All Intents V2 | ED CDC Firearm Assault V1 | ED CDC Firearm Intentional Self-Harm V1 | ED CDC Firearm Unintentional / Undetermined V1 | Not binned into any ED firearm injury-related syndrome |
| --- | --- | --- | --- | --- | --- | --- |
| **Assault** | 1945 | 1794<br>(92.2%) | 355<br>(18.3%) | 6<br>(0.3%) | 1234<br>(63.4%) | 151<br>(7.8%) |
| **Unintentional (self, other, or unknown shooter)** | 509 | 472<br>(92.7%) | 9<br>(1.8%) | 3<br>(0.59%) | 378<br>(74.3%) | 37<br>(7.3%) |
| **Intentional Self-Harm / Suicide** | 171 | 136<br>(79.5%) | 3<br>(1.8%) | 38<br>(22.2%) | 77<br>(45.0%) | 35<br>(20.4%) |
| **Undetermined**<br>**(self, other, or**<br>**unknown**<br>**shooter)** | 776 | 721<br>(92.9%) | 126<br>(16.2%) | 6<br>(0.77%) | 482<br>(62.1%) | 55<br>(7.1%) |

<sup>a</sup> An ED visit can be binned into 0,1 or more syndromes.

### EMS-ED linkage results

We then examined the linkage between the firearm injury events in our EMS analysis with ED data with the goal of comparing firearm injury intent across the two data sources. Of the 8854 EMS encounters included in our analysis, 6054 had a disposition of ‘*Patient Treated, Transported by this EMS Unit’* and, of those, 5558 had a destination type of ‘*Hospital-Emergency Department’*, ‘*Freestanding Emergency Department’* or ‘*Missing*’, and 5353 of those were transported to an ED in NC DETECT (i.e., all civilian, acute care EDs in NC), based on a review of the free text destination name. We were able to link 3401 (63.5%) of these EMS encounters to a corresponding ED visit (Figure 2).

**Fig 1.**
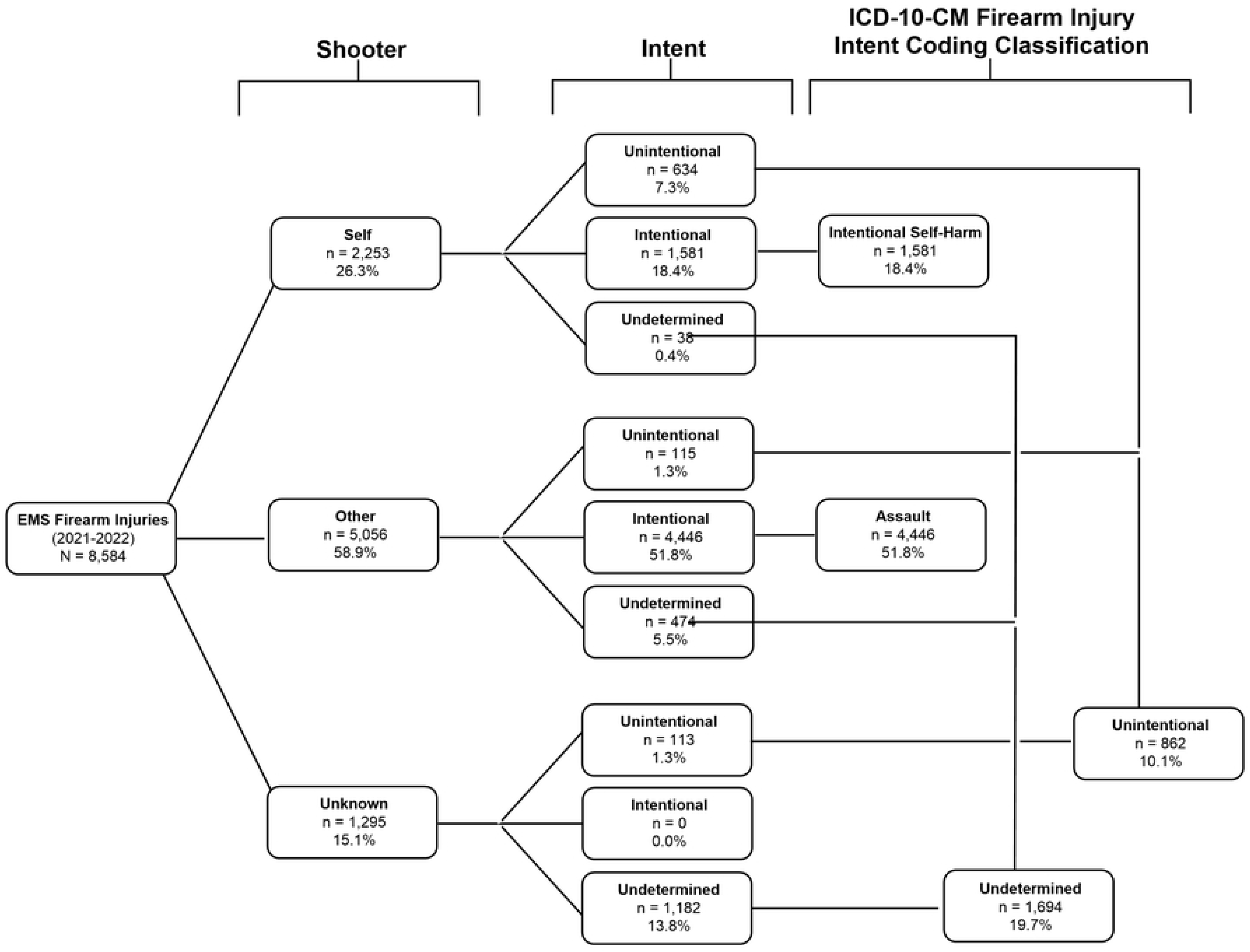
EMS Firearm Injuries by shooter and intent

**Fig 2.**
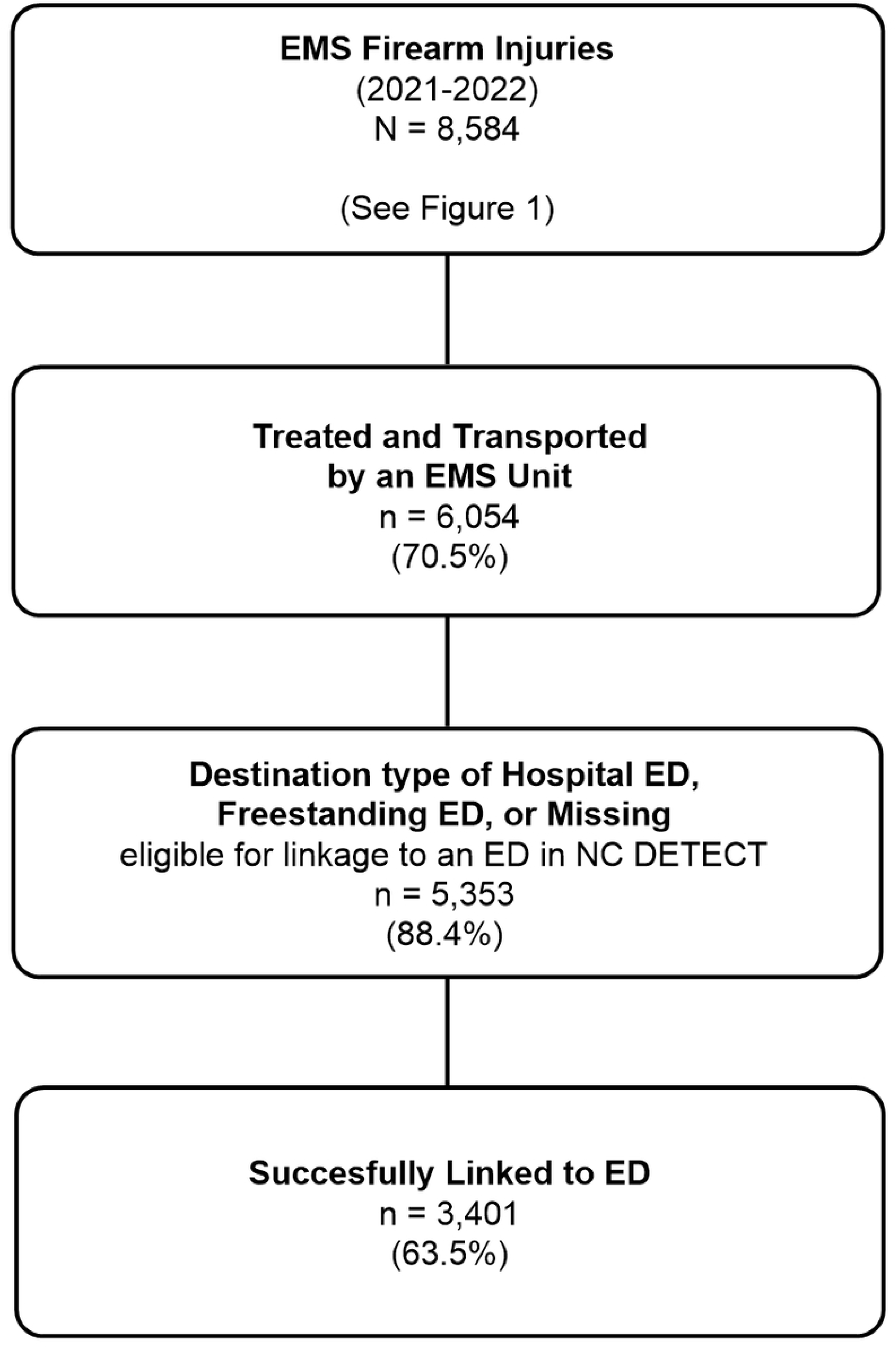
EMS-ED linkage results

For the EMS records that we successfully linked to an ED visit record (n = 3401), only 18.3% (n=355) of the assaults we identified in the EMS data (n=1945) were binned into the CDC Firearm Assault v1 definition, and only 22.2% (n=38) of the intentional self-harm suicide encounters (n=171) were binned into the CDC Firearm Intentional Self-Harm v1 definition. These findings illustrate the challenges of accurate intent coding in the ED setting and the potential impact of the CMS coding guidance that requires injury intent to default to unintentional if the intent is not identified in the physician documentation [17]. They also suggest that EMS encounters may serve as a better proximate for understanding intent for firearm injury than ED data, given the current coding guidance. EMS encounters identified as firearm injuries that were not identified as firearm injuries in the ED data often had trauma-related chief complaints and/or had diagnosis codes for penetrating injuries without an external mechanism firearm ICD-10-CM code.

While this study did not include a comprehensive review of EMS encounters transported to an ED that were not successfully linked to an ED visit record, our review of missed linkages revealed that missing or inaccurate dates of birth represented a significant cause of missed linkages. For serious events such as firearm injury, a reliable date of birth may be challenging for EMS to record at the scene.

### ED disposition for EMS-ED linked encounters

As shown in Table 7, 1721 (50.6%) of the ED visits linked to an EMS encounter had an ED disposition of discharged home, with 26.8% (n=913) admitted; 230 (6.8%) had a disposition of died in the ED. Of those who died in the ED, 76 (33%) were identified in the EMS encounter as intentional self-harm, and 103 (44.8%) as assault. These data indicate the overall severity of firearm injury and the lethality of firearms when used with intent to inflict injury. Only 2 (0.8%) deaths in the ED were for EMS identified unintentional firearm injuries.

**Table 7.** ED Disposition for EMS-ED linkage.

| **ED Disposition for EMS-ED Linked Data** | **Count (%)** |
| --- | --- |
| Discharged to home or self-care (routine discharge) | 1721 (50.6%) |
| Admitted to Hospital | 913 (26.8%) |
| Died | 230 (6.8%) |
| Transferred/discharged to another short-term general hospital | 249 (6.5%) |
| Transferred/discharged to another type of care setting | 65 (1.9%) |
| Transferred/discharged to prison or jail | 54 (1.6%) |
| Left AMA / Discontinued Care | 36 (1.1%) |
| Unknown/Missing | 133 (3.9%) |
| Total | 3401 (100%) |

## Limitations

The analysis presented in this report has several limitations. When manually coding intent, we relied on the EMS narrative but if the reporting of the incident intent to or by EMS was inaccurate, then the resulting intent coding may be inaccurate. For example, if the patient stated that they were shot in a drive-by shooting we coded this EMS encounter as an assault, but the patient may have unintentionally shot themselves and reported it as a drive-by shooting. In addition, our descriptive analysis included both nonfatal and fatal EMS encounters. While this reflects the comprehensive EMS data for firearm injury, certain findings may be different when analyzing nonfatal and fatal EMS firearm injury encounters separately. Finally, most of the EMS encounters in this analysis were reviewed by only one reviewer; intent classifications may have been assigned differently by a different reviewer.

## Conclusions

Timely and accurate injury cause codes are crucial for understanding the nature and scope of firearm-related injuries. In this study, we manually assigned intent to 8584 eligible NC EMS encounters from 2021 and 2022 to better understand the landscape of EMS-related firearm injury calls and to assess the accuracy of firearm injury coding in the ED setting. Manual coding of the data for shooter and intent revealed comparable results to those obtained from examining EMS assigned injury cause code classifications, when available. Of the 63.5% encounters (n = 3401) that could be linked to ED visit data, 1,945 of those were coded in the EMS data as assaults (57.2%). However, only 18.3% (n = 355) of those encounters were classified as assaults based on the ED CDC Assault v1 definition. Similarly, of the 171 encounters coded as intentional self-harm suicide encounters in the EMS, only 22.2% (n = 38) were binned to the CDC Firearm Intentional Self-Harm v1 definition. The ED data likely reflects current CMS coding guidance that requires injury intent to default to unintentional if the intent is not identified in the physician documentation [17].

The outcomes of our study reveal that EMS encounter data may be useful for obtaining more granular data from the scene of the injury incident. Such data might be important for developing tailored approaches that reflect regional and demographic differences in firearms injuries. In contrast, the lack of such detailed information in the ED visit data may limit such interventions. For example, if an encounter is the result of an assault, but it is coded as unintentional or undetermined, a patient may not be flagged for post-discharge follow-up that is directed towards supporting victims of assault. Combined, the results of this work illustrate the importance of accurate intent coding for firearms injuries and provides a foundation for the discussion of better coding guidance in the ED setting.

## Data Availability

NC DETECT emergency department visit data are available through a Data Use Agreement (DUA) with the NC Division of Public Health. More information is available at https://ncdetect.org/data-requests-for-applied-public-health-research/. NC emergency medical services (EMS) data are available from the NC Office of EMS through a DUA. for details on the DUA process.

## Acknowledgements

The North Carolina Disease Event Tracking and Epidemiologic Collection Tool (NC DETECT) is an advanced, statewide public health surveillance system. NC DETECT is supported by the North Carolina Division of Public Health through a federal Public Health Emergency Preparedness Grant and is managed through a collaboration between NC DPH and the University of North Carolina at Chapel Hill Department of Emergency Medicine’s Carolina Center for Health Informatics. The findings and conclusions in this publication are those of the author(s) and do not necessarily represent the views of the North Carolina Department of Health and Human Services, Division of Public Health.

The NC OEMS and the NC EMS Data System supports state, regional and local EMS and healthcare related service delivery from a patient care, resource allocation, and regulatory perspective. This manuscript was not prepared in collaboration with investigators of the NC EMS Data System and does not necessarily reflect the opinions or views of the NC OEMS, ESO or the study sites participating in the NC EMS Data System.

The authors also wish to thank Matthew Kerber for his work on the EMS-ED linkage components of this study.

## Notes

### Competing Interest Statement

The authors have declared no competing interest.

### Funding Statement

Yes

### Author Declarations

This work was reviewed by the IRB of the University of North Carolina at Chapel Hill and determined to be exempt. UNC IRB #21-3074.

